# Genome-wide meta-analysis, fine-mapping, and integrative prioritization identify new Alzheimer’s disease risk genes

**DOI:** 10.1101/2020.01.22.20018424

**Authors:** Jeremy Schwartzentruber, Sarah Cooper, Jimmy Z Liu, Inigo Barrio-Hernandez, Erica Bello, Natsuhiko Kumasaka, Toby Johnson, Karol Estrada, Daniel J. Gaffney, Pedro Beltrao, Andrew Bassett

## Abstract

Genome-wide association studies (GWAS) have discovered numerous genomic loci associated with Alzheimer’s disease (AD), yet the causal genes and variants remain incompletely identified. We performed an updated genome-wide AD meta-analysis, which identified 37 risk loci, including novel associations near genes *CCDC6, TSPAN14, NCK2*, and *SPRED2*. Using three SNP-level fine-mapping methods, we identified 21 SNPs with greater than 50% probability each of being causally involved in AD risk, and others strongly suggested by functional annotation. We followed this with colocalisation analyses across 109 gene expression quantitative trait loci (eQTL) datasets, and prioritization of genes using protein interaction networks and tissue-specific expression. Combining this information into a quantitative score, we find that evidence converges on likely causal genes, including the above four genes, and those at previously discovered AD loci including *BIN1, APH1B, PTK2B, PILRA*, and *CASS4*.

## Introduction

Genome-wide association studies (GWAS) for family history of disease, known as GWAS-by-proxy (GWAX), are a powerful method for performing genetic discovery in large, unselected cohort biobanks, particularly for age-related diseases^1^. Recent meta-analyses have combined GWAS of diagnosed late-onset Alzheimer’s disease (AD) with GWAX for family history of AD in the UK Biobank^2,3^, and reported a total of 12 novel disease-associated genomic loci. However, the causal genetic variants and genes which influence AD risk at these and previously discovered loci have only been clearly identified in a few cases. Discovering causal variants has led to deeper insight into molecular mechanisms of multiple diseases, including obesity^4^, schizophrenia^5^, and inflammatory bowel disease^6^. For AD, known causal variants include the *ε*4 haplotype in *APOE*, the strongest genetic risk factor for late-onset AD, and a common nonsynonymous variant that strongly alters splicing of *CD33* exon 2^7^. In addition, likely causal rare nonsynonymous variants have been discovered in *TREM2*^*8*^, *PLCG2* and *ABI3*^*9*^. These findings have strengthened support for a causal role of microglial activation in AD.

Although non-synonymous variants are highly enriched in trait associations, most human trait-associated variants do not alter protein-coding sequence and are thought to mediate their effects via altered gene expression, which is likely to occur in a cell type-dependent manner. A growing number of studies have mapped genetic variants affecting gene expression traits, known as expression quantitative trait loci (eQTLs), in diverse tissues or sorted cell types^10,11^. While it has become common to integrate GWAS results with eQTLs, this is often limited to a small number of datasets thought to be relevant.

To identify putative causal genetic variants for AD, we performed a meta-analysis of GWAX in the UK Biobank with the latest GWAS for diagnosed AD^12^, followed by fine-mapping using three alternative methods. Notably, this updated GWAS tested more genetic variants than the Lambert et al. study^13^ used in the meta-analyses by Jansen^2,3^ and Marioni^2^ (11.5 vs. 7.1 million). The increased power from our meta-analysis enabled us to discover four additional AD risk loci at genome-wide significance, and the higher density genotype imputation identified new candidate causal variants at both novel and established risk loci. We also performed statistical colocalisation analyses with a broad collection of eQTL datasets, including a recent study on primary microglia^14^, to identify candidate genes mediating risk at AD loci. We find that multiple lines of evidence, including colocalisation, tissue- or cell type-specific expression and prioritization using information propagation in gene networks, converge on a set of likely causal AD genes.

## Results

### Meta-analysis discovers 37 loci associated with Alzheimer’s disease risk

We performed a GWAX in the UK Biobank for family history of AD, based on 53,042 unique individuals who were either diagnosed with AD or who reported at least one first-degree relative (parent or sibling) having dementia, and 355,900 controls. This identified 13 risk loci at genome-wide significance (p < 5×10^−8^), 10 of which have been reported previously. Three novel loci were located near genes *NCK2, PRL*, and *FAM135B*. Notably, PRL has been reported as a CSF biomarker of AD^15^. We next did a fixed-effects meta-analysis of these GWAX results with the Kunkle et al. stage 1 GWAS meta-analysis of 21,982 cases with diagnosed AD and 41,944 controls^12^, across 10,687,126 overlapping variants (Figure 1). This revealed 34 AD risk loci (p < 5×10^−8^), 22 of which were reported in the Kunkle et al. study, while 8 others were reported in either Jansen et al.^3^ or Marioni et al^2^. Four loci were novel, located near genes *NCK2, TSPAN14, SPRED2*, and *CCDC6*. Notably, the *PRL* and *FAM135B* regions showed no evidence of association in Kunkle et al. (p > 0.1), and hence were not significant in meta-analysis. Three additional loci were found at suggestive significance (p < 5×10^−7^) in the meta-analysis, near genes *IKZF1, TSPOAP1*, and *TMEM163*. We included these loci in our follow-up analyses, for a total of 37 loci (Figure 1, Supplementary Table 1).

**Figure 1:**
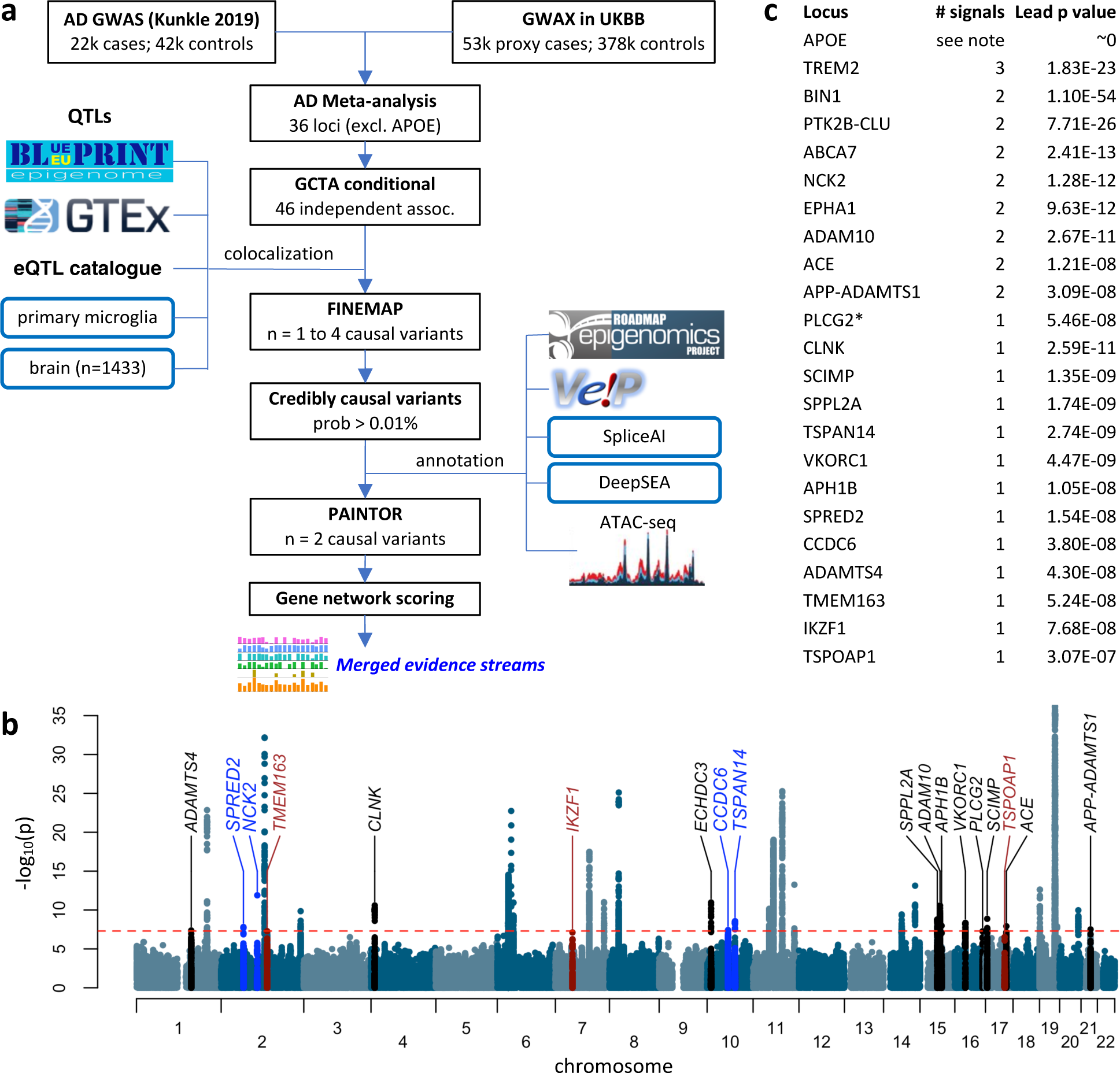
Analysis overview. (a) Summary of AD meta-analysis and data processing steps.Manhattan plot of the meta-analysis of GWAS for diagnosed AD and our GWAX in UK Biobank. Novel genome-wide significant loci are labelled in blue, sub-threshold loci in red, and recently discovered loci^2,3,12^ replicated in our analysis in black. (c) The number of independent signals at each locus which is either recently discovered or which has more than one signal. * The *PLCG2* locus was significant (p < 5×10^−8^) when including Kunkle stage 3 SNPs. Conditional analyses were not done at *APOE* due to the strength of the signal.

**Table 1:** Top candidate variants. A selected list of the most likely causal variants across loci, based on a combination SNP fine-mapping probabilities and annotations. Column ‘SNP prob’ indicates the mean fine-mapping probability for the SNP; the SpliceAI score is the maximum splicing probability for donor gain/loss or acceptor gain/loss, with nonzero values highly enriched for splicing effects; the DeepSEA functional significance score represents the significance above expectation for chromatin feature changes, as well as evolutionary conservation, with lower values more significant. References for specific SNPs are shown^2,7,9,14,40,41,60–69^.

| Gene | SNP | P value | Beta | Effect allele | Allele freq | SNP prob | SpliceAI | DeepSEA | Note | Refs |
| --- | --- | --- | --- | --- | --- | --- | --- | --- | --- | --- |
| ADAMTS4 | rs2070902 | 1.64E-06 | -0.0522 | T | 0.2580 | 0.384 | 0.107 | 0.140 | Intronic in candidate gene FCER1G, with predicted splicing change. |  |
| ADAMTS4 | rs4575098 | 4.30E-08 | 0.0609 | A | 0.2350 | 0.339 |  | 0.033 | 3' UTR of ADAMTS4, open chromatin | 2 |
| SPRED2 | rs268120 | 2.08E-08 | 0.0612 | A | 0.2502 | 0.556 |  | 0.033 | Strong DNase peak, predicted by DeepSEA to decrease. |  |
| NCK2 | rs143080277 | 1.28E-12 | -0.5205 | T | 0.9957 | 1.000 |  | 0.086 | Enhancer (Roadmap) |  |
| BIN1 | rs6733839 | 1.10E-54 | 0.1556 | T | 0.3915 | 0.998 |  | 0.027 | Microglia ATAC peak. DeepSEA predicts decreased DNaseHS. | 14,40 |
| INPP5D | rs10933431 | 1.41E-10 | 0.0770 | C | 0.7817 | 0.833 |  | 0.022 |  | 60 |
| PILRA | rs1859788 | 3.28E-18 | -0.0897 | A | 0.3206 | 0.601 | 0.008 | 0.041 | Known PILRA missense G78R. | 41 |
| ECHDC3 | rs7920721 | 1.08E-11 | -0.0669 | A | 0.6195 | 0.641 |  | 0.026 | DNase peak. DeepSEA predicts changed binding of USF, Max, Myc. | 61,62 |
| TSPAN14 | rs1870137 | 2.93E-09 | -0.0702 | C | 0.2056 | 0.097 |  | 0.007 | Top DeepSEA variant, predicting decreased binding of HNF4, FOXA1, SP1. |  |
| TSPAN14 | rs1870138 | 4.51E-09 | -0.0693 | A | 0.2057 | 0.068 |  | 0.004 | Highlighted in text; predicted loss of TAL1 binding. |  |
| SORL1 | rs11218343 | 5.59E-14 | 0.1864 | T | 0.9630 | 1.000 |  | 0.209 |  | 63 |
| SORL1 | rs2298813 | 1.52E-04 | 0.0850 | A | 0.0470 | 0.451 | 0.054 | 0.003 | Secondary assoc. Missense; also top DeepSEA variant. |  |
| APH1B | rs117618017 | 1.05E-08 | 0.0850 | T | 0.1395 | 0.895 | 0.007 | 0.019 | Highlighted in text; missense Thr27Ile. | 64 |
| PLCG2 | rs12444183 | 5.46E-08 | -0.0533 | A | 0.3830 | 0.686 |  | 0.220 | Near promoter of ncRNA AC099524.1, with strong microglia coloc. | 2 |
| PLCG2 | rs72824905 | 6.35E-06 | 0.2703 | C | 0.9924 | 0.492 | 0.018 | 0.006 | Secondary association; known missense Pro522Arg. Top DeepSEA score. | 9 |
| TSPOAP1 | rs2632516 | 3.12E-07 | -0.0493 | C | 0.4426 | 0.412 |  | 0.126 | Overlaps ncRNA containing mir-142, important for hematopoietic dev't. | 62,65 |
| TSPOAP1 | rs2526377 | 8.45E-07 | 0.0477 | A | 0.5579 | 0.169 |  | 0.006 | Top DeepSEA variant (decreased DNaseHS) in microglial ATAC peak. | 66 |
| ACE | rs4311 | 1.21E-08 | -0.0543 | T | 0.4704 | 0.490 | 0.126 | 0.053 | Strong predicted splicing change. | 67,68 |
| ACE | rs3730025 | 2.58E-07 | -0.1994 | A | 0.9828 | 0.416 | 0.002 | 0.021 | Secondary association; low-freq missense Tyr244Cys. |  |
| ABCA7 | rs12151021 | 2.41E-13 | 0.0773 | A | 0.3258 | 0.713 | 0.013 | 0.312 | Lead ABCA7 variant. |  |
| ABCA7 | rs4147918 | 7.63E-07 | 0.1201 | A | 0.9587 | 0.552 | 0.071 | 0.045 | Secondary association; missense Gln905Arg; predicted splicing change. | 69 |
| CD33 | rs12459419 | 2.02E-08 | -0.0576 | T | 0.3256 | 0.662 | 0.001 | 0.070 | Known missense Ala14Val; strong splicing QTL. | 7 |
| CASS4 | rs6014724 | 1.07E-10 | 0.1094 | A | 0.9122 | 0.548 |  | 0.083 | Lead CASS4 variant. |  |
| CASS4 | rs17462136 | 1.01E-09 | -0.1038 | C | 0.0872 | 0.067 |  | 0.001 | 5' UTR of CASS4; global top DeepSEA variant predicting decreased TF binding. |  |
| ADAMTS1 | rs2830489 | 3.09E-08 | -0.0590 | T | 0.2749 | 0.718 |  | 0.077 | Lead variant near ADAMTS1. |  |

Next, we applied stepwise conditioning using GCTA^16^, with linkage disequilibrium (LD) determined from UK Biobank samples, to identify independent signals at the discovered loci. We excluded the *APOE* locus from conditional analyses and fine-mapping, because the strength of the association made these analyses unreliable (see methods). Apart from *APOE*, 9 loci had two independent signals, while the *TREM2* locus had three signals (Figure 1c). Interestingly, a number of the loci discovered recently^2,3,12^ had multiple signals; specifically, *NCK2, EPHA1, ADAM10, ACE*, and *APP-ADAMTS1*. To extract insight from both the new and established AD GWAS discoveries, we performed comprehensive colocalisation, annotation, fine-mapping and network analyses to identify causal genes and variants (Fig 1a).

### Colocalisation between AD risk loci and gene expression traits

To identify genes whose expression may be altered by risk variants, we performed statistical colocalisation^17^ between each of 36 risk loci (excluding *APOE*) and a set of 109 eQTL datasets representing a wide variety of tissues, cell types and conditions (Figure 2, Supplementary Table 2). The eQTL datasets include a study of primary microglia from 93 brain surgery donors^14^, a meta-analysis of 1433 brain cortex samples^18^, as well as 49 tissues from the genotype-tissue expression project (GTEx) final release^10^, and 57 eQTL datasets uniformly reprocessed as part of the eQTL catalogue^11^. The latter include multiple studies in tissues of potential relevance to AD, such as brain, as well as sorted blood immune cell types under different stimulation conditions^19–35^.

**Figure 2:**
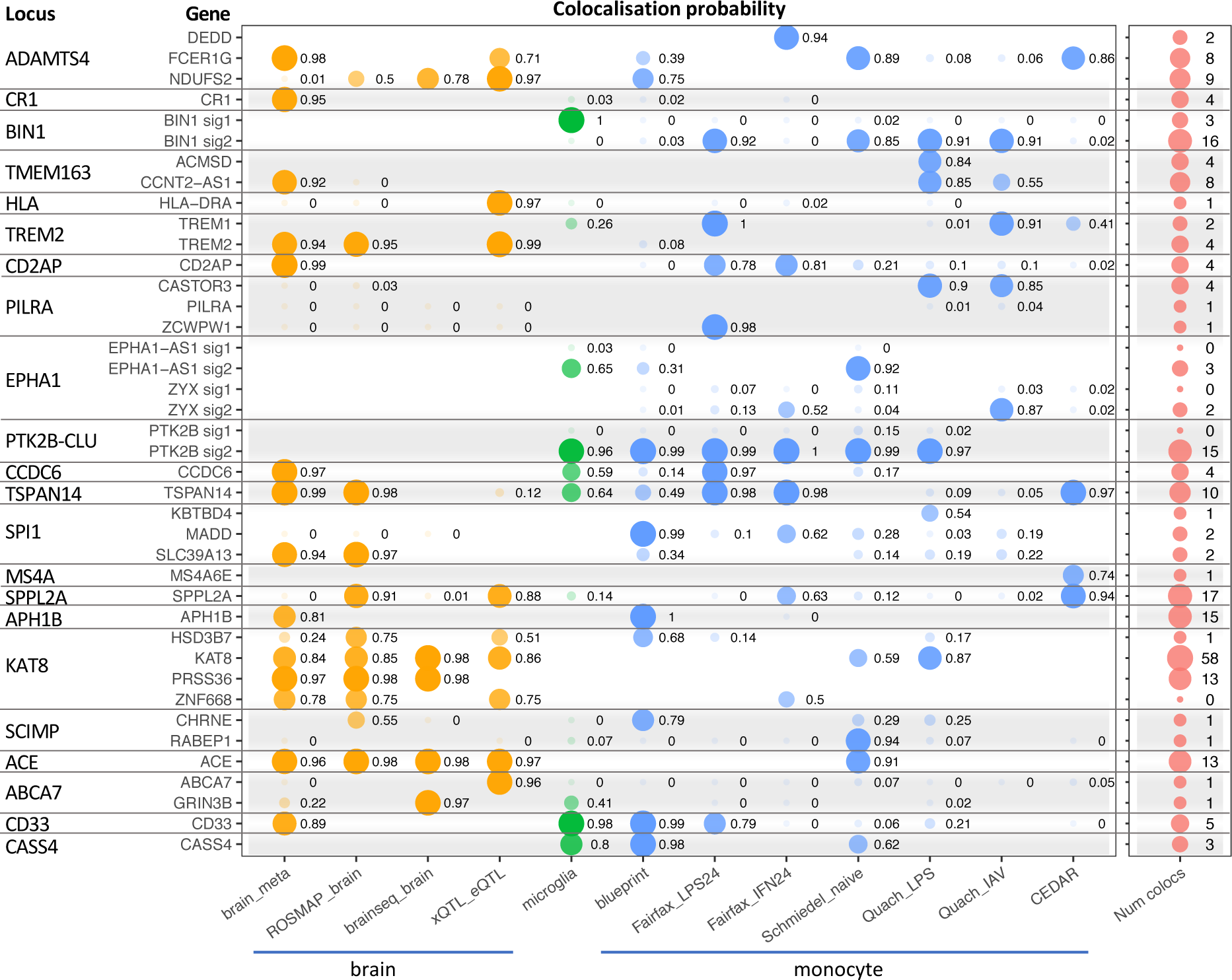
Colocalisation with eQTLs. For genes with the top overall colocalisation scores across AD risk loci, the colocalisation probability (H4) is shown for selected brain, microglia, and monocyte eQTL datasets. For three loci with multiple signals (*BIN1, EPHA1, PTK2B-CLU*), scores are shown separately for the conditionally independent signals. The last column shows, for each gene, the number of eQTL datasets with a colocalisation probability above 0.8 (Supplementary Tables 2-3).

Some studies using colocalisation have suggested that there is relatively limited overlap between GWAS associations and gene expression QTLs above that expected by chance^6,36^. A possible reason is that colocalisation analyses can suffer from a lack of sensitivity to detect shared causal variants between traits, which could occur for a number of reasons. First, when a locus has multiple causal variants, and not all causal effects are shared between a pair of studies (e.g. GWAS and eQTL study), colocalisation may not be detected^17^. Second, differences in LD patterns in a pair of studies can reduce the likelihood of a positive colocalisation. Third, relatively low power in either study can further reduce the colocalisation probability. To mitigate the first effect, we performed colocalisations separately for each conditionally independent AD signal, to model the case where not all causal variants are shared, as well as for the main AD signal at each locus. Problems relating to power and LD mismatch are partially mitigated by our use of a large number of the most highly-powered eQTL datasets currently available.

Across the 36 loci, we found 391 colocalisations with at least 80% probability of a shared causal variant between AD and eQTL, representing 80 distinct genes at 27 loci (Supplementary Tables 3-4). The genes implicated by colocalisation include many which have alternative lines of evidence for roles in AD, such as *PTK2B*^*37,38*^, *BIN1*^*39,40*^, *PILRA*^*41*^, *CD33*^*42,43*^, and *TREM2*^*44,45*^, as well as novel candidates including *FCER1G, TSPAN14, APH1B*, and *ACE*. However, the presence of multiple genes with colocalisation evidence within individual loci suggests that additional lines of evidence are important for prioritizing relevant genes.

Due to the large number of tissue datasets and colocalisation tests performed, we hypothesized that it would be important to upweight colocalisations in “relevant” tissues, as well as to accumulate colocalisation information across datasets. We therefore developed a weighted score which accumulates towards a maximum of 1.0 (inspired by scoring systems in STRING^46^ and Open Targets^47^), with higher weight on colocalisations in microglia, brain, and immune cell types than in other tissues (see methods). We compared this with a score obtained by taking the maximum colocalisation probability across all datasets.

To evaluate the ability of these scores to identify relevant AD genes, we considered the top 40 genes at our AD loci that were prioritized via other lines of evidence (namely, expression, distance, coding variant changes, and network score, described further below; genes in Supplementary Table 5). The top 80 genes by weighted coloc score retrieved 18 of the 40 genes. Surprisingly, the top 80 genes by maximum coloc score retrieved 20 genes (16 overlapping; Supplementary Figure 1), and we therefore used this score in subsequent gene prioritization. These results suggest that strong colocalisations in tissues not thought to be relevant can still be informative for identifying likely causal genes, and that with our approach, upweighting “relevant” QTL datasets showed no benefit for gene prioritization.

### Fine-mapping identifies credibly causal variants

Confirming the causal genes underlying AD risk will ultimately require experiments to identify the molecular mechanisms by which gene function is altered. Such experiments must be motivated by strong hypotheses regarding potentially causal variants and their possible effects. We sought to identify candidate causal variants using three distinct fine-mapping methods. First, we used the WTCCC Bayesian fine-mapping method^48^, which assumes a single causal variant, on each conditionally independent signal. Second, we used FINEMAP^49^ at each locus, specifying the number of independent signals determined by GCTA as the maximum number of causal variants per locus. Third, we used PAINTOR^50^, a method which estimates enrichments in functional genomic annotations to obtain a posterior probability of causality for each variant based on its annotations, and which also can account for multiple causal variants. For computational feasibility with PAINTOR, we only considered variants with at least 0.01% causal probability as determined by FINEMAP.

We used 43 annotations individually as input to PAINTOR (Supplementary Table 6); these included ATAC-seq peaks from primary microglia^51^ or iPSC-derived macrophages^52^, DNase peaks from cell type groups in the Roadmap Epigenomics project^53^, variant consequence annotations^54^ and evolutionary conservation^55^ (Figure 3b). We also used scores from DeepSEA^56^ and SpliceAI^57^, deep-learning methods that predict the effects of variants on transcription factor binding or splicing. Missense mutations were the most enriched annotation, with a 19.2-fold increased odds of being causal SNPs, but they comprised only 1% of input SNPs. Blood or immune DNase hypersensitivity peaks merged from 24 Roadmap Epigenomics tissues provided the highest model likelihood, as these peaks covered 16% of SNPs, despite a lower 6.4-fold enrichment. Variants with a nonzero score from the SpliceAI method, which predicts changes to gene splicing, were also highly enriched (9.3-fold), while variants with top DeepSEA scores were more modestly enriched.

**Figure 3:**
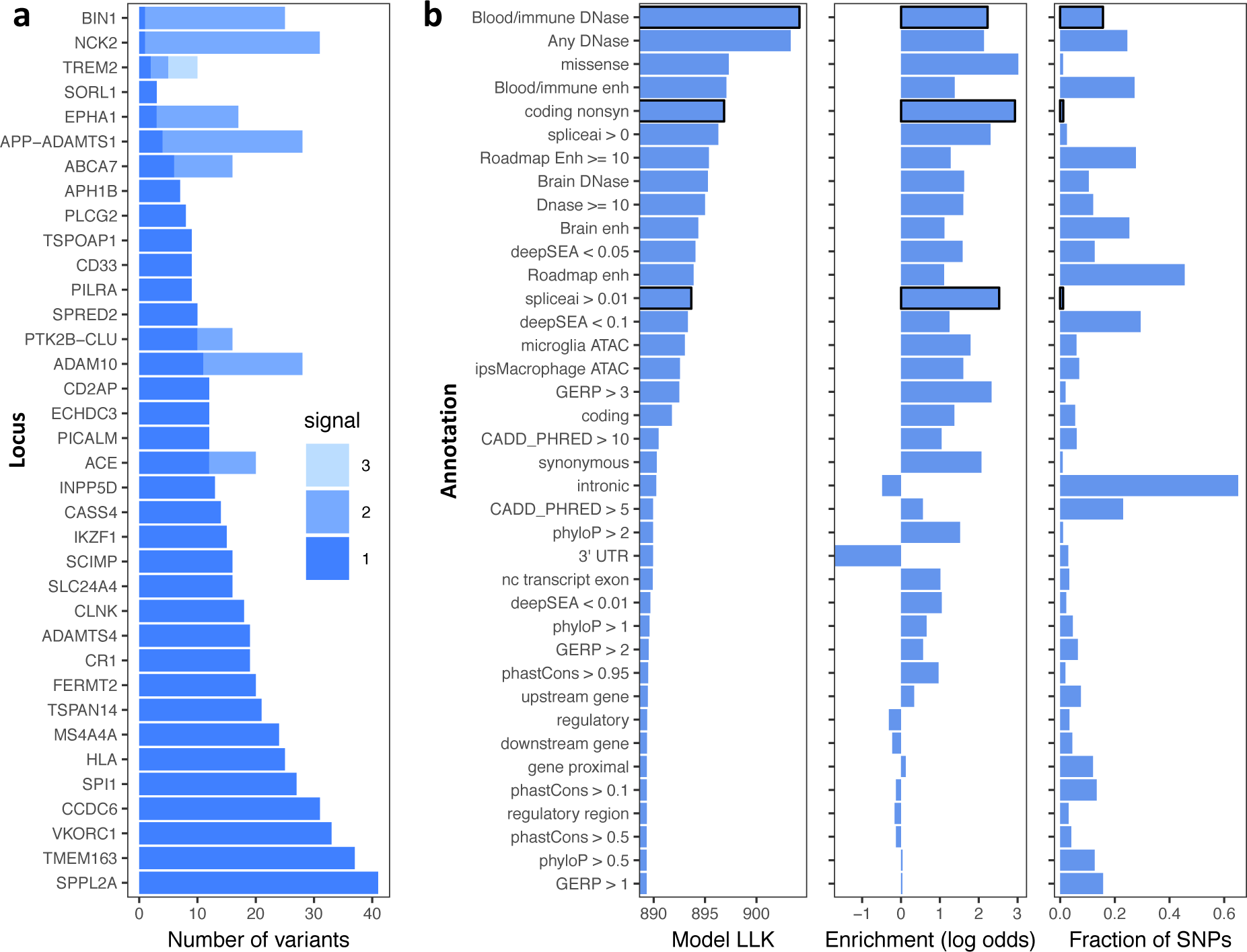
Fine-mapping summary. (a) Number of variants with mean causal probability > 1% for each independent signal. Variant counts for independent signals are shown in different shades. (b) PAINTOR outputs, showing (left) log-likelihood (LLK) of model for each individual annotation; (middle) log-odds enrichments for individual genomic annotations determined by PAINTOR; (right) fraction of SNPs which are in each annotation (among those selected by FINEMAP probability > 0.01%). Annotations selected for the final model are shown with a black border.

We next built a multi-annotation model in PAINTOR (v3.1) following a stepwise selection procedure, which identified a minimal but informative set of three annotations: blood and immune DNase, nonsynonymous coding variants, and variants with SpliceAI score greater than 0.01. We used probabilities from this PAINTOR model, and computed the mean causal probability per variant across the three fine-mapping methods.

There were 21 variants with a mean causal probability above 50% across the fine-mapping methods, and 79 further variants with probabilities from 10 - 50% (Table 1 and Supplementary Table 7). These include SNPs near established AD risk genes, such as rs6733839 ∼20 kb upstream of *BIN1*, which has recently been shown to alter a microglial MEF2C binding site^14^ and to regulate *BIN1* expression specifically in microglia^40^. High-confidence variants also include a well-known missense SNP in *PILRA*^*41*^, and a splice-altering missense SNP in *CD33*^*7*^. Missense SNP rs4147918 in *ABCA7* had 55% causal probability, and *ABCA7* harbored 5 further missense SNPs with FINEMAP probabilities greater than 0.01%, at varying allele frequencies. Notably, rs4147918 as well as 6 other variants within *ABCA7*, including the lead SNP rs12151021, had positive SpliceAI scores for predicted changes to gene splicing. This is consistent with reports of a burden of deleterious variants at *ABCA7* associated with AD^58^, as well as potential changes to splicing caused by intronic variable tandem repeats^59^.

A number of newly identified AD risk genes had high-confidence fine-mapped variants. These include the *NCK2* rare intronic SNP rs143080277 (>99% probability, MAF 0.4%), *APH1B* missense SNP rs117618017 (90% probability), rs2830489 at the *APP-ADAMTS1* locus (72% probability), rs61182333 intronic in *SCIMP* (61% probability), and rs268120 intronic in *SPRED2* (56% probability).

Annotation-based fine-mapping highlighted a number of candidate causal variants, which were not always the highest probability SNPs at the locus (Figure 4). Within *TSPAN14*, rs1870137 and rs1870138 reside within a DNase hypersensitivity peak found broadly across tissues, which is also an ATAC peak in microglia. Of these, rs1870138 lies at the centre of a ChIP-seq peak for binding of multiple transcription factors, including FOS/JUN and GATA1, and is within a FOS/JUN motif, albeit at a relatively low information content position. However, the alt allele rs1870138-G alters an invariant position of a binding motif for *TAL1*, a gene highly expressed in microglia, and which is a binding partner for GATA1. The AD risk allele, rs1870138-G, is also associated with increased monocyte count^70^ and increased risk for inflammatory bowel disease^71^, and in both cases is among the top associated variants. Notably, the AD signal in the region colocalises with both an eQTL and a splicing QTL for *TSPAN14* in multiple datasets, and rs1870138-G associates with higher *TSPAN14* expression in brain and in microglia, but with lower expression in some GTEx tissues.

**Figure 4:**
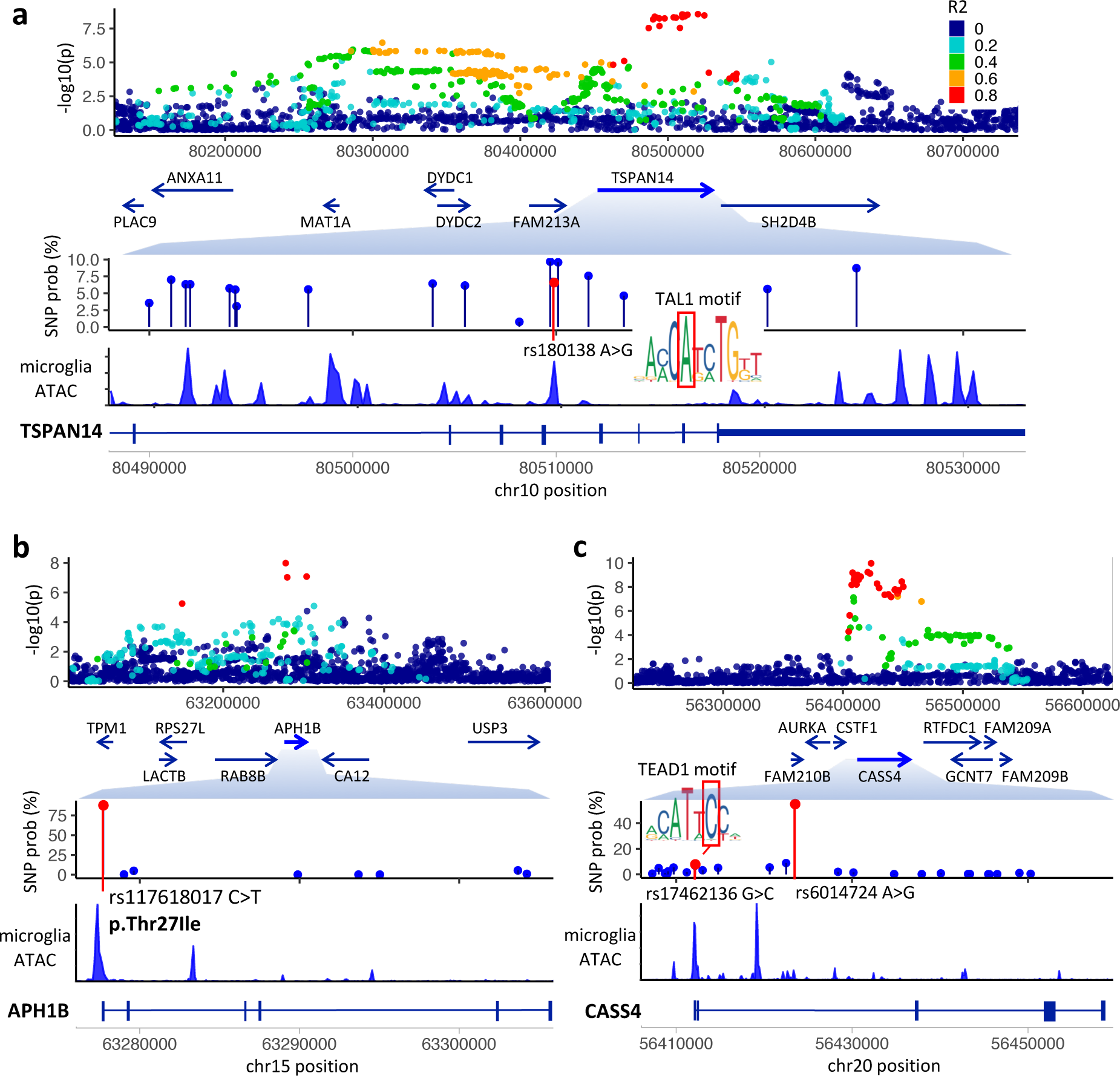
Fine-mapped variants. (a) SNP rs1870138 in an intron of *TSPAN14* disrupts an invariant position of a TAL1 motif. (b) Missense SNP rs117618017 in exon 1 of *APH1B*. (c) SNP rs17462136 in the 5’ UTR of *CASS4* introduces a TEAD1 motif.

Missense SNP rs117618017 in exon 1 of *APH1B* (T27I) is the likely single causal variant at its locus, with fine-mapping probability of 90% (Figure 4b). APH1B is a component of the gamma-secretase complex, other members of which (*PSEN1, PSEN2*) have rare variants associated with early-onset AD^72^. Interestingly, the AD signal colocalises with an *APH1B* eQTL in monocytes, neutrophils and T-cells, as well as numerous GTEx tissues, and the rs117618017-T allele associates with higher AD risk and higher *APH1B* expression across datasets. rs117618017-T introduces a motif for transcriptional regulator YY1, and is predicted by DeepSEA to increase YY1 binding in multiple ENCODE cell lines. Therefore, it is an open question whether AD risk is mediated by altered APH1B protein structure or altered gene expression.

Finally, the AD association on chromosome 20 colocalises with an eQTL for *CASS4* in Blueprint monocytes and in GTEx whole blood, as well as in fibroblasts. While lead SNP rs6014724 (55% probability), intronic in *CASS4*, shows no evidence of transcription factor (TF) binding in ENCODE data, rs17462136 (7% probability) lies in a region of dense TF binding in the 5’ UTR of *CASS4* (Figure 4c). The nucleotide position is highly conserved (GERP score 3.46), overlaps an ATAC peak in microglia, and the rs17462136-C allele introduces a TEAD1 binding motif, making it the strongest functional candidate SNP. In addition, rs17462136 is more strongly associated with *CASS4* expression in multiple eQTL datasets than is rs6014724.

### Network evidence prioritizes genes within and beyond GWAS loci

As a further line of evidence, we developed a method that leverages gene network connectivity to prioritize genes at individual loci. We first constructed a gene interaction network combining information from the STRING, IntAct and BioGRID databases. Next, we nominated candidate genes at each AD locus (Supplementary Table 8), based on a mix of our other evidence sources as well as literature reports, and used these as seed genes similar to the approach used in the priority index for drug discovery^73^. For each locus in turn, we used as input all seed genes except those at the locus, and propagated information through the network with the page rank algorithm (see methods). The “networkScore” for a gene thus represent the degree to which the gene is supported by its interaction with top AD candidate genes across all other loci, unbiased by any locus-specific features.

Across AD loci, likely candidate genes were highly enriched for having high network-based gene scores (Wilcoxon rank sum test, p = 5×10^−14^; Supplementary Figure 2). Notably, at our four novel AD loci, the nearest gene (*NCK2, TSPAN14, SPRED2, CCDC6*) was the highest-scoring gene among those within 200 kb, and in each case was one of the top two highest-scoring genes within 500 kb. Many established or recently discovered AD genes were also the top gene within 500 kb by network score, including *ACE, CASS4, CD2AP, PICALM, PLCG2, PTK2B*, and *TREM2*. At the *SLC24A4* locus, *RIN3* was strongly supported, whereas *SLC24A4* was not, in line with evidence from deleterious rare variants that *RIN3* may be causal^12^. At the *ECHDC3* locus, both *USP6NL* and *CELF2* had high network scores, while at the *EPHA1* locus, *ZYX* was the top scoring gene. Interestingly, AD candidate genes *ABCA7* and *CR1* had only modest network scores, suggesting a need to integrate across independent lines of evidence to prioritize genes.

Genes highly ranked by network propagation also include many outside of genome-wide significant AD loci (Supplementary Table 9). Consistent with their involvement in AD, genes ranked in the top 500 by network score tended to have SNPs with lower p values nearby (within 10 kb) than did remaining genes (Wilcoxon rank sum test, p = 7×10^−7^), suggesting that there remain numerous AD loci to be discovered with larger GWAS sample sizes. Top network-ranked genes include *LILRB2* (nearby SNP p = 9.8×10^−6^), a leukocyte immunoglobulin-like receptor that recognizes multiple HLA alleles, and which may also be involved in amyloid-beta fibril growth^74^; *ABCA1* (SNP p = 4×10^−6^), involved in phospholipid transfer to apolipoproteins and previously associated with AD^75^; *SREBF1* (SNP p = 2×10^−6^), required for lipid homeostasis; *AGRN* (SNP p = 4×10^−6^), involved in synapse formation in mature hippocampal neurons; and *CD19* (SNP p = 1×10^−5^), an antigen coreceptor on B-lymphocytes. Overall, genes with high network ranks were strongly enriched in biological processes and pathways that have previously been associated with AD, including clathrin-mediated endocytosis, activation of immune response, phagocytosis, Ephrin signaling, and complement activation (Supplementary Table 10).

### Integrative gene prioritization from five lines of evidence

We developed a comprehensive gene prioritization score, which incorporates quantitative information from multiple evidence sources, unlike binary indicator scores that have been used by others for GWAS gene prioritization^12,76^ (Figure 5, Supplementary Table 11). The five lines of evidence include gene distance to lead SNPs, expression level in either microglia or brain tissue, colocalisation score, network score, and the sum of fine-mapped probability for any coding SNPs within a gene (see methods for details).

**Figure 5:**
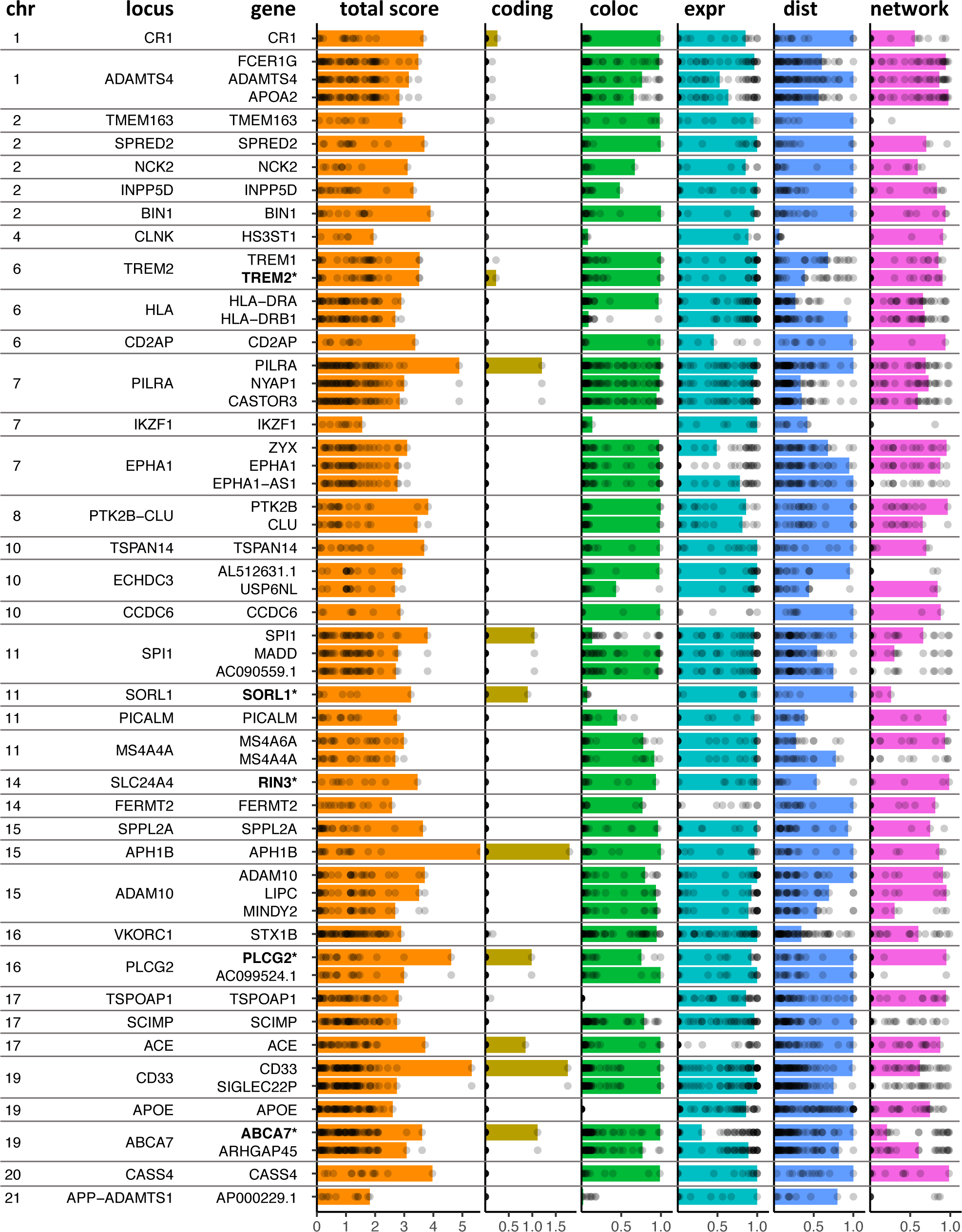
Gene evidence summary. The top gene at each locus is shown, as well as the next 16 top genes by total score. Score components for each gene are indicated by coloured bars, and points show the distribution of scores for all genes within 500 kb at the locus. Bold gene names are those with evidence of causality based on rare variants from other studies.

To incorporate gene distance, we used a function that decays from 1 to 0 with increasing log-scaled distance up to 500 kb (Supplementary Figure 3). Although it is clear that long-distance gene regulation can occur, recent evidence from both eQTLs^77^ and metabolite GWAS^78^ suggests that genomic distance from the association peak is a strong predictor of causal target genes. For gene expression, we considered either the expression level of genes in microglia or brain, or the specificity of each gene’s expression in these cells / tissues relative to all GTEx tissues. We found that specificity of expression in microglia or brain was at least as informative as the absolute level of expression in these tissues, and that incorporating expression level did not improve a score based solely on expression specificity (Supplementary Figure 4). Hence, we used only expression specificity in our overall prioritization.

The comprehensive prioritization score identified the majority of AD candidate genes previously suggested as causal (Figure 5), suggesting that our evidence sources complement each other. Indeed, although the network score is independent of locus-specific features, gene ranks based on network scores within each locus were highly correlated with gene ranks based on the other four evidence sources (Spearman rho=0.47; p < 2.2×10^−16^). Exemplifying the importance of integrating genetic evidence, *ABCA7, SORL1*, and *CR1* were top-ranked by overall score at their respective loci, despite having only moderate network-based scores, while *SORL1, PICALM* and *SPI1* were top-ranked despite having limited eQTL colocalisation evidence.

While our prioritisation further supports many established AD candidate genes, it also implicates novel genes. Among these are *FCER1G* at the *ADAMTS4* locus, a gene which negatively regulates immune cell responses^79^ and has been reported as a hub gene in microglial gene modules associated with neurodegeneration^80,81^. Another candidate is *ZYX* at the *EPHA1* locus, which receives a top network score, is highly expressed in microglia, and which was recently nominated as an AD risk gene based on chromatin interactions between the *ZYX* promoter and AD risk variants in a *ZYX* enhancer^82^. Finally, at the MS4A locus our prioritisation nominates *MS4A6A*, another gene implicated in AD risk based on chromatin interactions^82^, despite this gene being relatively distal (143 kb) from the AD association peak.

## Discussion

Identifying therapeutic targets for human diseases is a key goal of human genetics research, and is particularly important for neurodegenerative diseases such as AD, for which no disease-modifying therapies yet exist. However, identifying the causal genes and genetic variants from GWAS is challenging: association peaks span large genomic regions, and non-coding associations can act via regulation of distal genes. We approached this challenge for AD by performing the largest meta-analysis to date, followed by comprehensive fine-mapping, eQTL colocalisation, and quantitative gene prioritization.

Our meta-analysis identified four novel associations with genome-wide significance, near *NCK2, SPRED2, TSPAN14*, and *CCDC6*. Each of these genes was supported by both eQTL colocalisation and network ranking, and in each case was the nearest gene to the association peak. Indeed, when distance was excluded from the priority score, for 21 of the 37 loci the top prioritized gene was the nearest gene, and for a further 8 loci the top gene was within 100 kb. This is consistent with observations from eQTL studies that the majority of gene regulatory variants lie within 100 kb of their regulated genes^83^.

Despite the large number of eQTL datasets that we used, colocalisation of likely AD risk genes was sometimes found in only one or a few datasets; this was the case for *SPRED2* (TwinsUK LCL coloc probability 0.99), *RIN3* (GTEx frontal cortex probability 0.94), and *PILRA* (Fairfax LPS-2hr monocyte coloc probability 0.99). Many factors could account for dataset-specific colocalisations, such as biological differences in sample state, differences in LD match between the GWAS and eQTL datasets, and technical differences in the transcriptome annotations used for eQTL discovery. As a result, absence of colocalisation provides only weak evidence for lack of an effect in a given tissue type, whereas positive colocalisation provides strong support for a shared genetic effect. It is therefore useful to look broadly across eQTL studies for colocalisation, which will be facilitated by resources that simplify access to these datasets, such as the eQTL catalogue^11^.

Our gene prioritization incorporated multiple lines of genetic evidence to distinguish genes most likely to causally mediate AD risk. This analysis supported roles for many established AD genes, while also pointing to novel candidates. One of our most confidently prioritized genes was *APH1B*, encoding a gamma-secretase complex component involved in APP processing. *APH1B* harbors the likely causal missense variant rs117618017, yet also has strong colocalisation evidence that higher expression correlates with higher AD risk. One possibility is that impaired function of *APH1B* due to the missense variant leads to upregulation of *APH1B* transcription. This interpretation would be consistent with evidence from both mice^84^ and humans^85^ that loss of APH1B and gamma-secretase function leads to AD. Although APH1B loss may be associated with non-AD dementia^84,86^, effect sizes for rs117618017 on risk were similar both for diagnosed AD (Kunkle et al. OR 95% CI=[1.071, 1.127]) and dementia in UK Biobank (OR 95% CI=[1.064, 1.104]).

Among our novel associations, *TSPAN14* has a role in defining the localisation of *ADAM10*^*87*^, another recently discovered AD gene which is a key component of the gamma-secretase complex, and which could thus mediate AD risk via processing of amyloid precursor protein. However, ADAM10 also cleaves the microglia-associated protein TREM2 to generate its soluble ligand-binding domain^88^. Our fine-mapping showed that the risk SNP rs1870138 is also associated with higher risk for inflammatory bowel disease (IBD), an immune-mediated disease, and with higher monocyte count in UK Biobank individuals. Since *TSPAN14* is expressed more highly in immune cell types, including microglia, than in brain tissue, it is also plausible that AD risk is mediated by its effect on either immune cell count or activation. *SPRED2* is a negative regulator of ERK/MAPK signalling, and its loss in mice leads to increased macrophage activation and tissue inflammation^89^. The AD-associated SNPs in *SPRED2*, rs268134 and rs268120, are also associated with increased neutrophil percentage and decreased lymphocyte percentage in the UK Biobank.

Recently proposed AD candidate genes supported by our analyses include *RIN3, HS3ST1*, and *FCER1G*. As noted above, *FCER1G* is a negative regulator of immune cell responses^79^; *RIN3* interacts with both *BIN1* and *CD2AP* in the early endocytic pathway^90^; *HS3ST1* is involved in cellular uptake of tau^91^ and was recently been associated with AD in an independent Norwegian sample^61^.

In summary, our study reports fine-mapping SNP probabilities for 36 AD-associated regions, including 4 novel loci. By combining evidence from eQTL colocalisations across 109 datasets, functional annotations and gene network analysis, we generate a quantitative prioritization score and provide a comprehensive map of AD candidate genes. Our genetic findings highlight the presence of diverse mechanisms in AD pathogenesis, suggesting different possible entry points for interventions to treat AD or to reduce risk of the disease, and identify candidate targets for therapeutic development.

## Methods

Code for analyses described here can be found at github.com/jeremy37/AD_finemap.

### GWAS on family history of AD

Sample QC, variant QC and imputation was performed on all UK Biobank participants as described in Bycroft et al.^92^. After genotype imputation, 93,095,623 variants across 487,409 individuals were available for analysis. To exclude individuals of non-European ancestry, we first extracted the “White British” ancestry subset of participants as described in Bycroft et al. 2018. These individuals self-reported their ethnic background as “British” have similar genetic ancestry based on principal components (PC) analysis. To extract additional individuals of European ancestry, we followed a similar approach to Bycroft et al. and applied Aberrant^93^ on PCs 1v2, 3v4 and 5v6 across the individuals who self-reported as “Irish” or “Any other white background”. Pairs of first-degree relatives were identified using KING v2.0^94^. We applied KING to 147,522 UK Biobank individuals who had at least one relative identified in Bycroft et al. (UK Biobank Field 22021). For each first-degree relative pair, we prioritized AD cases and proxy-cases (see below) for inclusion, and otherwise excluded one of the pair at random. We also excluded variants with low imputation quality (INFO < 0.3) and/or those with minor allele frequencies below 0.0005, resulting in 25,647,815 variants available for analysis.

AD cases were extracted from UK Biobank self-report (field 20002), ICD10 diagnoses (fields 41202 and 41204) and ICD10 cause of death (fields 40001 and 40002) data. UK Biobank participants were asked whether they have a biological father, mother or sibling who suffered from Alzheimer’s disease/dementia (UK Biobank fields 20107, 20110 and 20111 respectively). We extracted all participants with at least one affected relative as proxy-cases. Participants who answered “Do not know” or “Prefer not to answer” were excluded from analyses. All remaining individuals were denoted as controls.

There were 3,046 AD cases, 52,791 AD proxy cases and 355,900 controls in the combined white British and white non-British cohorts. For association analyses, we lumped the true and proxy-cases together (53,042 unique affected individuals) and used the linear-mixed model implemented in BOLT-LMM^95^.

### AD meta-analysis

To enable meta-analysis combining the UK Biobank cohorts with external case-control studies, we first transformed the AD proxy BOLT-LMM summary statistics from the linear scale to a 1/0 log odds ratio:

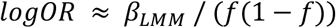

with standard error:

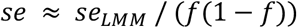

where β_LMM_ and se_LMM_ are the SNP effect sizes and standard errors respectively from BOLT-LMM, and *f* is the fraction of cases in the sample^96^. Since the affected individuals in our analysis include both true and proxy-cases, we then multiplied the transformed logORs and standard errors by 1.897 so that it approximates the logORs obtained from a true case/control study^1^.

We combined the transformed UK Biobank white British cohort, the transformed UK Biobank white non-British cohort and the Stage 1 summary statistics from Kunkle et al. using a fixed-effects (inverse variance weighted) meta-analysis across 10,687,126 overlapping variants. For display purposes (Supplementary Table 7), we used CrossMap^97^ to convert variant positions from GRCh37 to GRCh38.

### Conditional analysis and statistical fine-mapping

To run GCTA, we prepared plink input files with genotypes from 10,000 randomly sampled UK Biobank individuals at variants within +/- 5 Mb from each lead SNP. We excluded variants with INFO < 0.85, or which had a p-value from Cochran’s Q test for study heterogeneity < 0.001. We also excluded variants with allele frequency in UK Biobank below 0.1%, as LD estimates are unreliable at low allele counts. We ran GCTA --cojo-slct with a p-value threshold of 10^−5^ to identify secondary signals at each locus, and then retained only loci with a lead p-value below 5×10^−8^. For the HLA locus we used a GCTA p-value threshold of 5×10^−8^. We also retained the loci *TSPOAP1, IKZF1*, and *TMEM163* since they had p < 5×10^−8^ in an earlier version of our analysis. We excluded the APOE locus from conditional analysis and fine-mapping because the strength of association in the region would require a more perfect LD panel match to avoid spurious signals.

We then ran FINEMAP at each locus, with --n-causal-snps given as the number of independent SNPs determined by GCTA. For FINEMAP, we excluded variants with allele frequency below 0.2%, since we found that otherwise FINEMAP sometimes selected implausible causal variants, such as pairs of very weakly associated rare variants to explain a common variant signal. For loci with multiple signals, we also used GCTA --cojo-cond to condition on each independent SNP identified in the previous analysis, and retained SNPs within 500 kb of any conditionally independent SNP at the locus. To compute SNP causal probabilities based on GCTA conditional signals, we converted effect size (beta) and standard error values to approximate Bayes Factors (BF)^98^ using a prior of W=0.1 (in Wakefield notation), and used the WTCCC single-causal variant method^48^, probability = SNP BF / sum(all SNP BFs).

### Colocalisation with eQTLs

For eQTL colocalisation, we downloaded summary statistics for the eQTL datasets mentioned in the main text, as well as the xQTL dataset^19^ based on dorsolateral prefrontal cortex brain samples. QTL calling for primary microglia was performed with RASQUAL^99^ with the --no-posterior-update option. We determined eQTL genes at FDR 5% for each dataset in a uniform manner, first using Bonferroni correction of lead SNP nominal p values based on the number of variants within 500 kb of the gene, and using the Benjamini-Hochberg method to compute FDR. We matched variants between eQTL and GWAS based on chromosomal position; for datasets in GRCh38 coordinates, we first used CrossMap^97^ to convert back to GRCh37 coordinates. We used the coloc package^17^ with default priors to perform colocalisation tests between GWAS and eQTL signals where the lead variants were within 500 kb of each other, and passed to coloc all variants within 200 kb of each lead variant. We also performed colocalisation tests using GWAS p-values for each conditionally independent GWAS signal, obtained with GCTA as described above.

### Functional annotations

All functional annotations used were in GRCh37 coordinates, as was the AD meta-analysis. We used the Ensembl VEP online Web tool (www.ensembl.org/vep)^54^ to predict variant consequences, and to add selected annotations (Supplementary Table 6). We downloaded bed files based on imputed data for Roadmap Epigenomics DNase, histone peaks, and 25-state genome segmentations for 127 epigenomes^53^. We grouped these into groups “all”, “brain” (epigenomes 7, 9, 10, 53, 54, 67, 68, 69, 70, 71, 72, 73, 74, 81, 82, 125), and “blood & immune” (epigenomes 33, 34, 37, 38, 39, 40, 41, 42, 43, 44, 45, 47, 48, 62, 29, 30, 31, 32, 35, 36, 46, 50, 51, 116). For genome segmentations, we considered 9 states to represent enhancers: TxReg, TxEnh5, TxEnh3, TxEnhW, EnhA1, EnhA2, EnhAF, EnhW1, EnhW2. We used bedtools^100^ to determine overlaps, and counted the number of overlaps for each variant with peaks in the above groups. We downloaded FANTOM5^101^ permissive enhancer annotations from fantom.gsc.riken.jp/5/data. We downloaded pre-computed SpliceAI scores^57^ for variants within genes from github.com/Illumina/SpliceAI. We merged filtered whole-genome and exome scores together to obtain the most comprehensive predictions, and for each AD variant we annotated the maximum predicted score across splice donor gain, donor loss, acceptor gain, acceptor loss. We used the DeepSEA^56^ online tool (deepsea.princeton.edu) to annotate variants selected for functional fine-mapping with DeepSEA’s “functional significance” score. BigWig files with PhastCons, PhyloP and GERP RS scores in hg19 (GRCh37) coordinates were downloaded from UCSC. We downloaded microglial ATAC-seq based on the study by Gosselin et al.^51^, aligned reads to GRCh37 with bwa 0.7.15^102^, and called multisample peaks across all 15 datasets using MACS2^103^. We prepared bigWig files from alignments by using bedtools genomecov, followed by bedGraphToBigWig. To visualise microglia ATAC-seq tracks we adapted code from wiggleplotr^104^.

### Annotation-based fine-mapping

For fine-mapping with PAINTOR, we selected 3,207 variants which had (a) FINEMAP probability >= 0.01% based on the GCTA-identified number of causal variants at the locus, or (b) had FINEMAP probability >= 1% when run with either 1 or 2 causal variants, even if this was not the number identified by GCTA, or (c) were among the top 20 variants at the locus by FINEMAP probability. We defined binary annotations for input to PAINTOR based on the features described above, which included thresholding certain scores at multiple levels (e.g. CADD >= 5, 10, 20). For Roadmap DNase and enhancer annotations, we included a category based on whether a variant was in a peak or enhancer in >= 10 epigenomes. We next ran PAINTOR v3.1 once for each of the 43 annotations (Figure 2; Supplementary Table 6), allowing 2 causal variants per locus. (In addition to excluding *APOE*, we excluded the *CLNK* locus because PAINTOR failed to run when this locus was input.)

To build a multi-annotation model, we performed forward stepwise selection. We selected the best annotation by model log-likelihood (LLK), Blood & immune DNase, and then ran PAINTOR again for each combination of this annotation and the 42 remaining annotations. We continued adding an annotation at each iteration from among those top-ranked by model LLK until the model LLK improvement was less than 1. This occurred at iteration 4, and so we kept the first 3 annotations in the combined model. We computed the mean causal probability for each SNP as the mean of the 3 fine-mapping methods at loci with two or more signals, or as the mean of the FINEMAP and PAINTOR probabilities for loci with one signal, since FINEMAP gives approximately the same results as WTCCC fine-mapping for a single causal variant.

### Network analysis

For network analysis, we created a gene interaction network based on selecting all edges between protein-coding genes from systematic studies (>1000 interactions) in the IntAct^105^ and BioGRID databases^106^, as well as edges from the STRING database version 10.5^46^ with edge score > 0.75. This combined network included 18,055 genes and 540,421 edges. We identified 36 top candidate genes across AD loci (Supplementary Table 8) to use as seed genes, and assigned weight to these according to the -log10(p value) of the lead SNP at the locus. 33 of the candidate genes were present in the network, while 3 were not (*ECHDC3, TMEM163, SCIMP*). For each locus, we used all seed genes as input except those at the same locus, and propagated information through the network with the personalized PageRank algorithm^107^, included in the igraph R package^108^. We found that a gene’s resulting PageRank was highly correlated with its node degree, and this made PageRank itself less informative. We therefore compared the PageRank of each gene at the locus to the distribution of PageRanks obtained for the same gene in 1,000 iterations of network propagation, where the same number of seed genes were randomly selected. We computed the percentile of a gene’s true PageRank relative to the 1,000 network propagations with randomized inputs. To determine gene set enrichment, we used the top 1,000 genes by network rank as input to gProfiler^109^ with default settings, with the set of all genes ranked by the network as a background set.

### Gene expression

Gene expression values for all tissues were determined in units of transcripts per million (TPM). Both GTEx v8 and the eQTL catalogue provide tables of the median TPM expression across samples for each tissue and gene. For primary microglia we obtained a table of read counts per gene, computed using FeatureCounts 1.5.3 as described in^14^, from which we computed median TPM.

### Gene prioritization

The **combined score** for each gene is the sum of five scores:

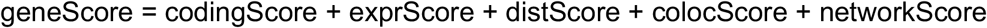

The **coding score** is twice the sum of the mean fine-mapping probability for missense or LoF variants in a gene.

The **expr(ession) score** is determined by first computing the percentile of expression (measured in TPM) for a gene in microglia relative to all GTEx tissues, and for GTEx brain (mean of brain tissues) relative to all other GTEx tissues. The score is then normalised to be in the range [0, 1], and rewards genes with expression percentile above the 50th:

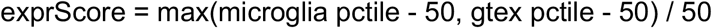

The **dist(ance) score** is defined as:

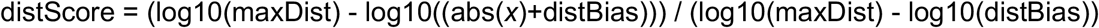

where *x* is the minimum distance of any portion of the gene footprint to the region defined by independent lead SNPs at a GWAS locus, maxDist is 500,000 and distBias is 6,355, chosen to give reasonable scores over the main range of interest of 0 - 200 kb (Supplementary Figure 3).

The **coloc score** used as part of the combined score is defined as the maximum value of the “H4” hypothesis probability output by the coloc R package. The weighted coloc score this was compared to was designed to accumulate evidence across datasets, prioritizing those most relevant, and is inspired by the Open Targets^47^ and StringDB^46^ evidence scoring systems. Each QTL dataset was assigned to one of the categories “relevant”, “possibly relevant”, and “not relevant”, which receive weights of 1.0, 0.8, and 0.5. Within each category, the score accumulated by ordering coloc H4 probabilities in descending order, and adding incrementally:

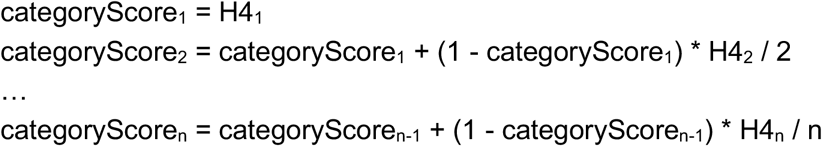

Within a category, then, a small number of strong colocs will receive a higher score than many weak colocs. The score was then accumulated across categories as:

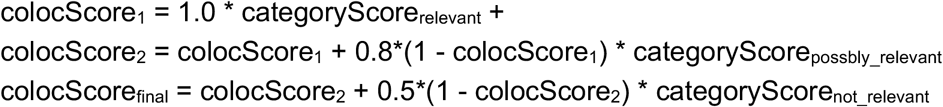

For example, if there were two “relevant” coloc H4 values of 0.4, and a “not relevant” H4 value of 0.9, the colocScore would be:

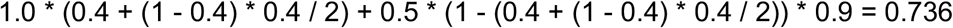

In contrast, with one “relevant” H4 value of 0.9 and two “not relevant” H4 values of 0.4, the colocScore would be:

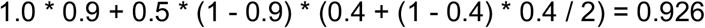

The **network score** is determined based on the page rank percentile for a gene relative to permutations:

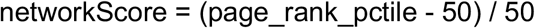

## Data Availability

Genome-wide summary statistics will be made available through the NHGRI-EBI GWAS catalog upon publication.

## Data availability

Summary statistics from the UK Biobank GWAX for AD and from the meta-analysis will be made available through the NHGRI-EBI GWAS Catalog: www.ebi.ac.uk/gwas/downloads/summary-statistics

## Acknowledgements

This work was funded by Open Targets (OTAR037). We thank Jeff Barrett for guidance during initiation of the project; Adam Young for sharing early access to human primary microglia data; and Kaur Alasoo for early access to the eQTL catalogue.

## Author contributions

JS planned and conducted the analyses, and wrote the paper. JL performed the GWAX and meta-analysis. SC and EB assisted with fine-mapping, variant and gene prioritization. IB and PB performed and supervised gene network analysis. NK performed microglia eQTL mapping. AB, TJ, DJG, and KE conceived and supervised the study.

## Supplementary Figures

**Supplementary Figure 1:**
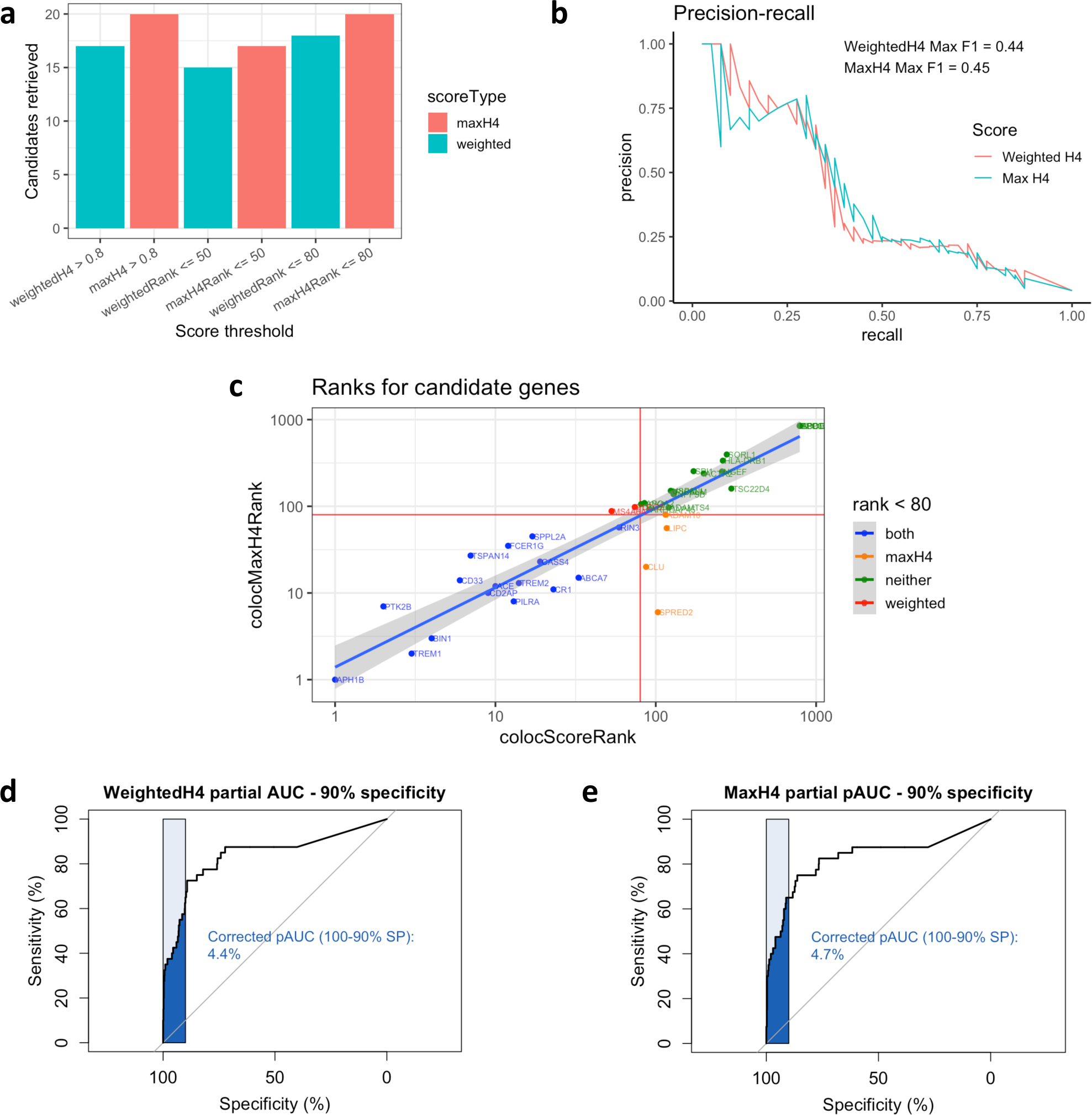
Maximum coloc probability (“maxH4”) outperformed a weighted coloc score that prioritises relevant cell types. We compared the performance of each score in identifying the top 40 AD candidate genes based on total score without coloc. (a) Barplot showing the number of candidate genes retrieved by either maxH4 or weighted coloc score at three score thresholds. In each case maxH4 retrieved more candidate genes. (b) Precision-recall curves, showing comparable F1 score between the two methods. (c) Scatter plot of gene ranks for top 40 genes relative to all genes, for weighted or maxH4 score. (d,e) Receiver operator characteristic partial area under the curve (AUC) at 90% specificity for (d) weighted coloc score and (e) maxH4 score. The maxH4 score showed slightly higher sensitivity at high specificity.

**Supplementary Figure 2:**
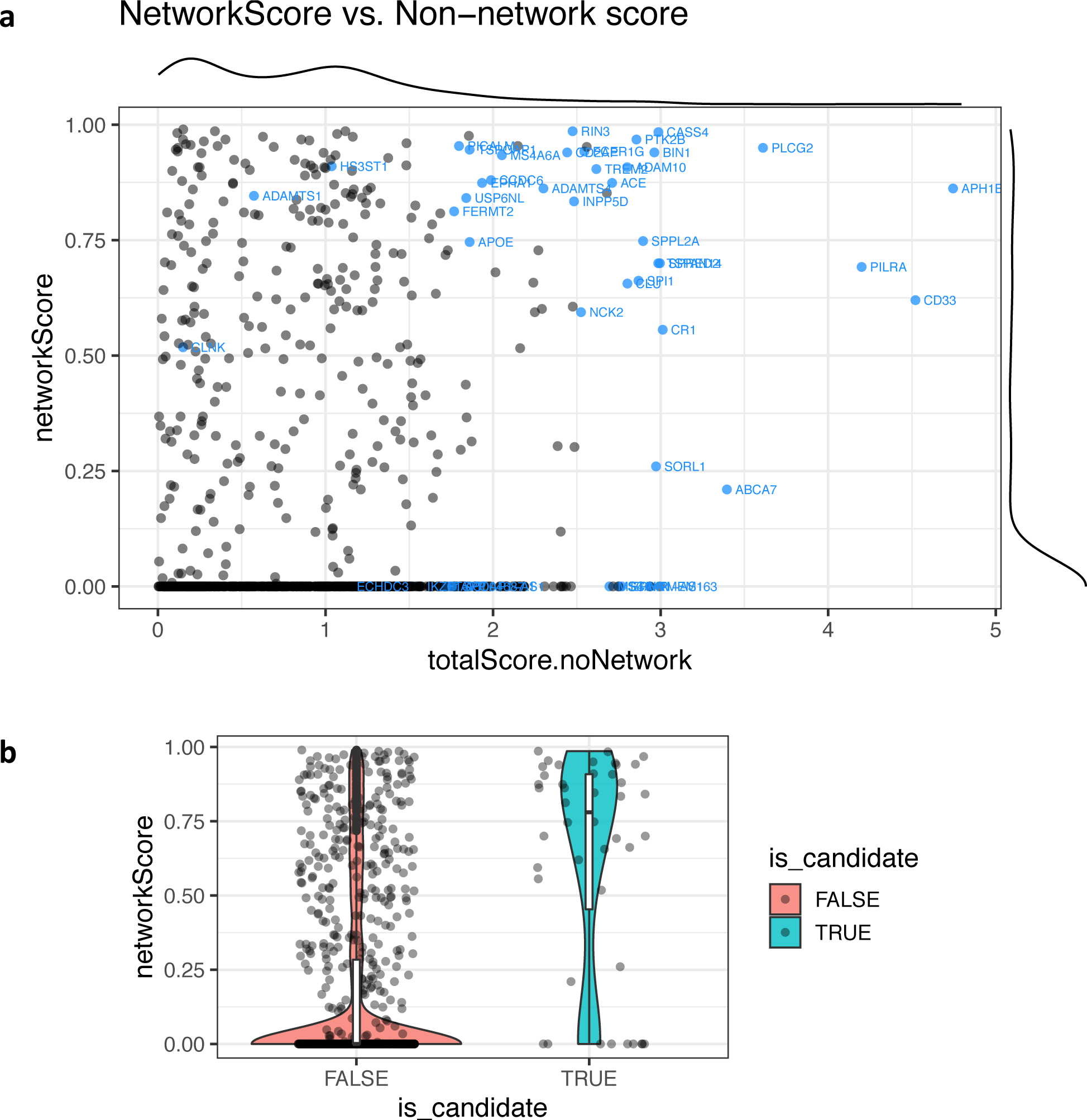
Network scores are higher for AD candidate genes. (a) Scatterplot of network score vs. the total score excluding network information (coloc + fine-mapping + expression + distance) for genes at 36 AD loci included in the main text. (b) Violin plot showing the distribution of scores for AD candidate genes (highlighted in part a) and all others. Note that genes not present in the network (*TMEM163, ECHDC3, SCIMP*) receive a score of zero by default.

**Supplementary Figure 3:**
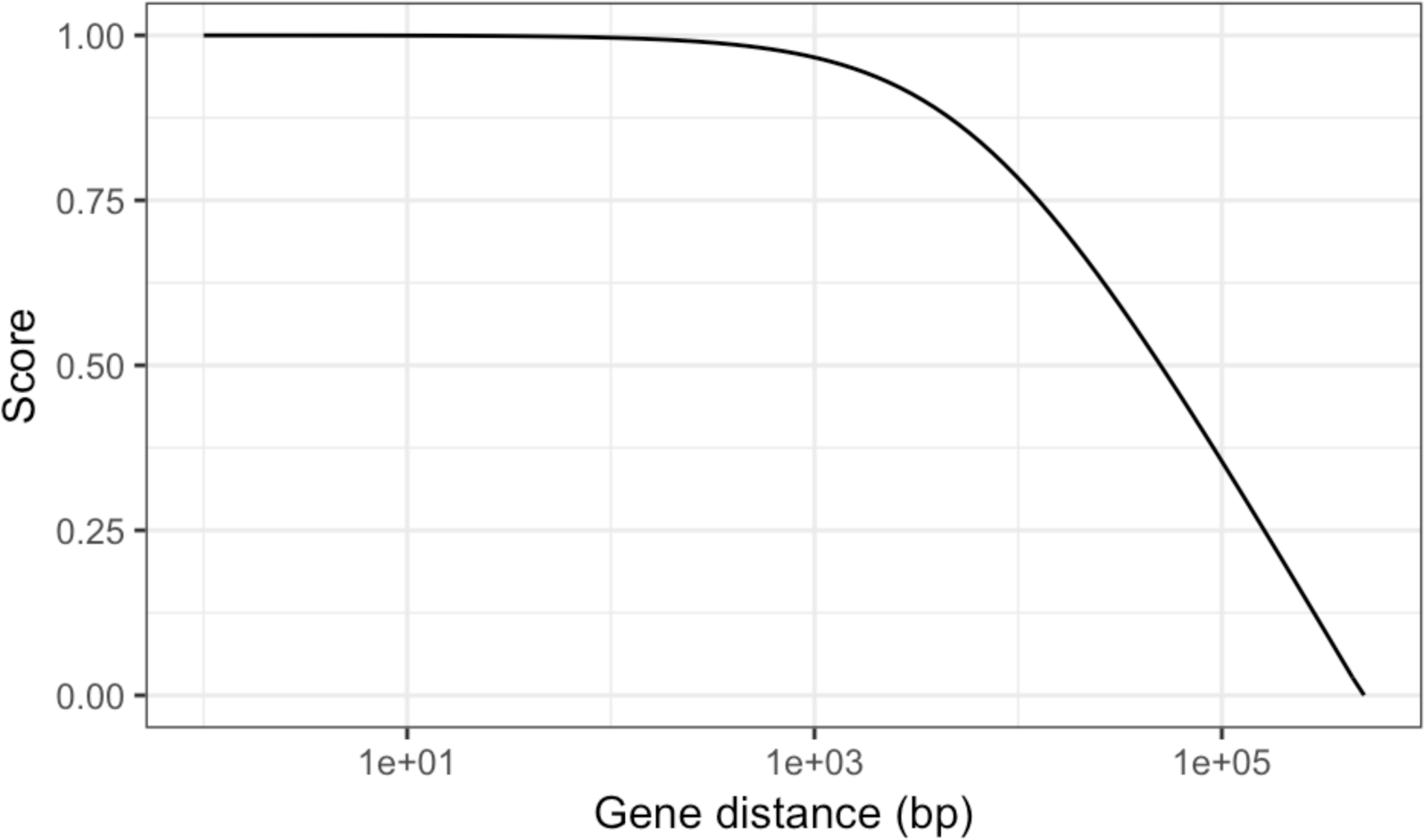
The gene distance score decreases with increasing log-scaled distance to the lead SNP.

**Supplementary Figure 4:**
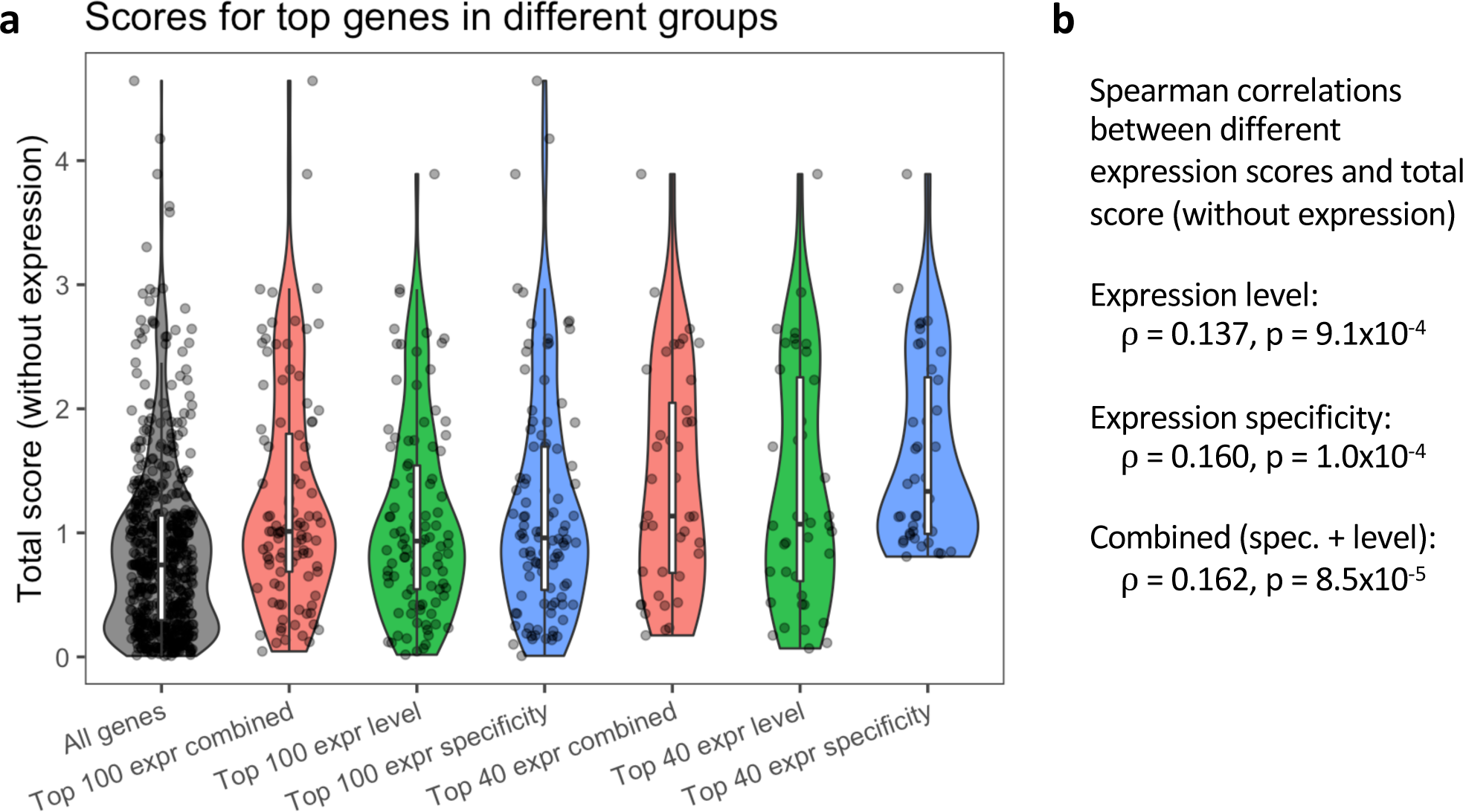
(a) Violin plots showing the distribution of the total gene score, excluding the expression component, in different groups of genes defined by variants of the expression score. Only protein-coding genes were included. For each of the expression scores, the top 100 or top 40 genes were selected. The expression scores compared were: expression level (log10(TPM) / 3); specificity of gene expression in brain or microglia relative to all GTEx tissues (described in methods); combined expression score (0.5 * exprLevelScore + 0.5 * specificityScore). (b) The score based on expression specificity in microglia or brain had slightly higher correlation with the total (non-expression) score than did the expression level score. A score combining both components showed similar correlation to that based on expression specificity alone.

